# Panel-agnostic tumour-only diagnosis of mismatch repair deficiency

**DOI:** 10.64898/2026.11.19.26368513

**Authors:** Diego Garcia-Lopez, Barbara Hernando, Blas Chaves-Urbano, Maria Escobar-Rey, David Gómez-Sánchez, Alice Cadiz, Alejandro Gallego, Marta Mendiola, Alberto Berjón, Eduardo Caleiras, Juan F. García, David Hardisson, Gemma Toledo, Andrés Redondo, María J. García, Geoff Macintyre

## Abstract

A diagnosis of mismatch repair deficiency (MMRd) can unlock immunotherapy treatment for a patient. However, existing sequencing-based detection methods require matched normal samples, panel-specific baselines or dedicated microsatellite loci, none of which are available to the tumour-only targeted panels that dominate routine clinical practice. Here we present captureMMRd, an open-source classifier that diagnoses MMRd using mutational signatures computed from the somatic variant calls any panel already produces. Applying a single locked model to 7,514 samples from 16 independent cohorts, spanning whole-exome, whole-genome, targeted panel and plasma cfDNA sequencing, captureMMRd achieved a pooled sensitivity of 94.36% and specificity of 99.02% against orthogonal MMR IHC and MSI-PCR testing, and outperformed microsatellite-based callers on panels with limited microsatellite content. We demonstrate that as few as 3 indels and 44 single-base substitutions are sufficient for reliable diagnosis, defining a minimum panel footprint that most clinically deployed assays already exceed. Our findings extend clinically actionable MMRd detection to the sequencing workflows of routine precision oncology, with the potential to broaden patient access to immunotherapy without new assays, baselines or matched normal samples.

## Introduction

Loss of mismatch repair proficiency in a tumour leads to the accumulation of replication errors, particularly at microsatellite loci, generating a microsatellite instability-high (MSI-H) phenotype and a high burden of tumour neoantigens. Accurate identification of MMRd/MSI-H tumours therefore has direct therapeutic relevance, most notably for selecting patients who may benefit from immune checkpoint blockade^1–3^, for Lynch syndrome screening^4,5^, and for prognostic and predictive information in several solid tumours^6,7^. This has made MMRd/MSI-H testing an essential component of modern precision oncology, extending beyond colorectal and endometrial cancer into pan-cancer diagnostic workflows^8^.

Clinical assessment of MMRd rests on immunohistochemistry (IHC) for loss of expression of the mismatch repair proteins MLH1, PMS2, MSH2 and MSH6, and/or PCR-based testing of microsatellite instability across a small set of markers^8–10^. These assays are well established and remain the clinical standard in many settings, but each captures only part of the biology of MMRd. IHC provides protein-level evidence of MMR pathway disruption yet can be affected by heterogeneous staining, retained expression of non-functional proteins and interpretative variability, while MSI-PCR directly measures microsatellite instability but interrogates a limited number of loci, with performance that varies across tumour types and sample contexts^11–13^.

Next-generation sequencing (NGS) offers an opportunity to integrate MMRd assessment into the molecular profiling many patients already receive. Sequencing-based approaches can simultaneously evaluate MSI, tumour mutational burden (TMB), mutational patterns and pathogenic alterations in MMR genes, providing a more comprehensive readout than single-modality testing^14–16^. These approaches span whole-genome (WGS) and whole-exome (WES) sequencing, through to comprehensive targeted panels, and MSI status is now reported by clinically implemented assays such as MSK-IMPACT and FoundationOne CDx as part of broader genomic profiling^17–19^. Yet each of these readouts is an imperfect proxy for MMRd. Classification based on MMR pathogenic variants can miss tumours driven by MLH1 promoter methylation, non-coding or regulatory alterations, or other epigenetic mechanisms not routinely captured by standard clinical panels^20–22^. TMB is enriched in MMRd tumours but is not specific to MMRd, as POLE/POLD1-deficient tumours can show extremely high TMB without MMRd^23,24^.

However, current sequencing-based MSI/MMRd detection remains anchored to the assay it was built for. Most methods infer instability by reading dedicated microsatellite loci, requiring matched normal sequencing^25,26^, a panel-specific baseline of microsatellite-stable samples^27,28^, models pre-trained on specific platforms^29^ or on assay-specific sequencing reads^30^, or proprietary microsatellite content designed into commercial panels^19,31,32^; even recent reference-free approaches require sufficient evaluable microsatellite loci on the panel^33^, with performance that varies with assay design and tumour type^30,34,35^. Mutation-based alternatives have so far been limited to single-feature heuristics, such as homopolymer indel counts calibrated against IHC in colorectal cancer within a single institutional assay^36^. The mutational consequences of MMRd are nonetheless directly visible in the somatic variant calls every panel already produces, and formal mutational-signature analysis capturing the full MMRd repertoire (SBS6, SBS14, SBS15, SBS20, SBS21, SBS26, SBS44, ID1, ID2 and ID7)^37–43^ has not been developed into a validated pan-cancer diagnostic. Yet most routine clinical NGS is performed using tumour-only capture sequencing, often to reduce cost, tissue requirements and turnaround time^44–47^, on panels that vary substantially in size, genomic content and microsatellite coverage, and that are processed through heterogeneous local bioinformatic pipelines. As a result, laboratories running tumour-only panels, particularly smaller or custom designs, have no validated route to MMRd assessment: they lack the matched normals, the baseline cohorts and the microsatellite content that existing methods assume. There is therefore a critical unmet need for an open, pan-cancer, panel-agnostic method that infers MMRd status directly from the somatic variant calls every targeted panel already captures, independent of microsatellite coverage, baselines or matched normal samples.

Here, we present captureMMRd, an open-source, capture-based classifier that reads MMRd-associated SBS and indel mutational signatures from routine tumour-only variant calls (**Figure 1a**). We train the model on tumour–normal WES data with orthogonal MSI-PCR reference testing and validate it, locked and without retraining, across 16 independent cohorts spanning tumour–normal WES, WGS and targeted panels, tumour-only institutional and custom panels, and plasma cfDNA. We benchmark captureMMRd against existing tumour-only MSI callers, assess its robustness to sample preservation method, and derive the minimum input for a reliable diagnosis (as few as 3 indels and 44 single-base substitutions) translating into a minimum panel footprint that most clinically deployed assays already exceed. By operating on the output sequencing workflows already generate, captureMMRd extends clinically actionable MMRd detection to the heterogeneous tumour-only panels used in routine precision oncology.

**Figure 1:**
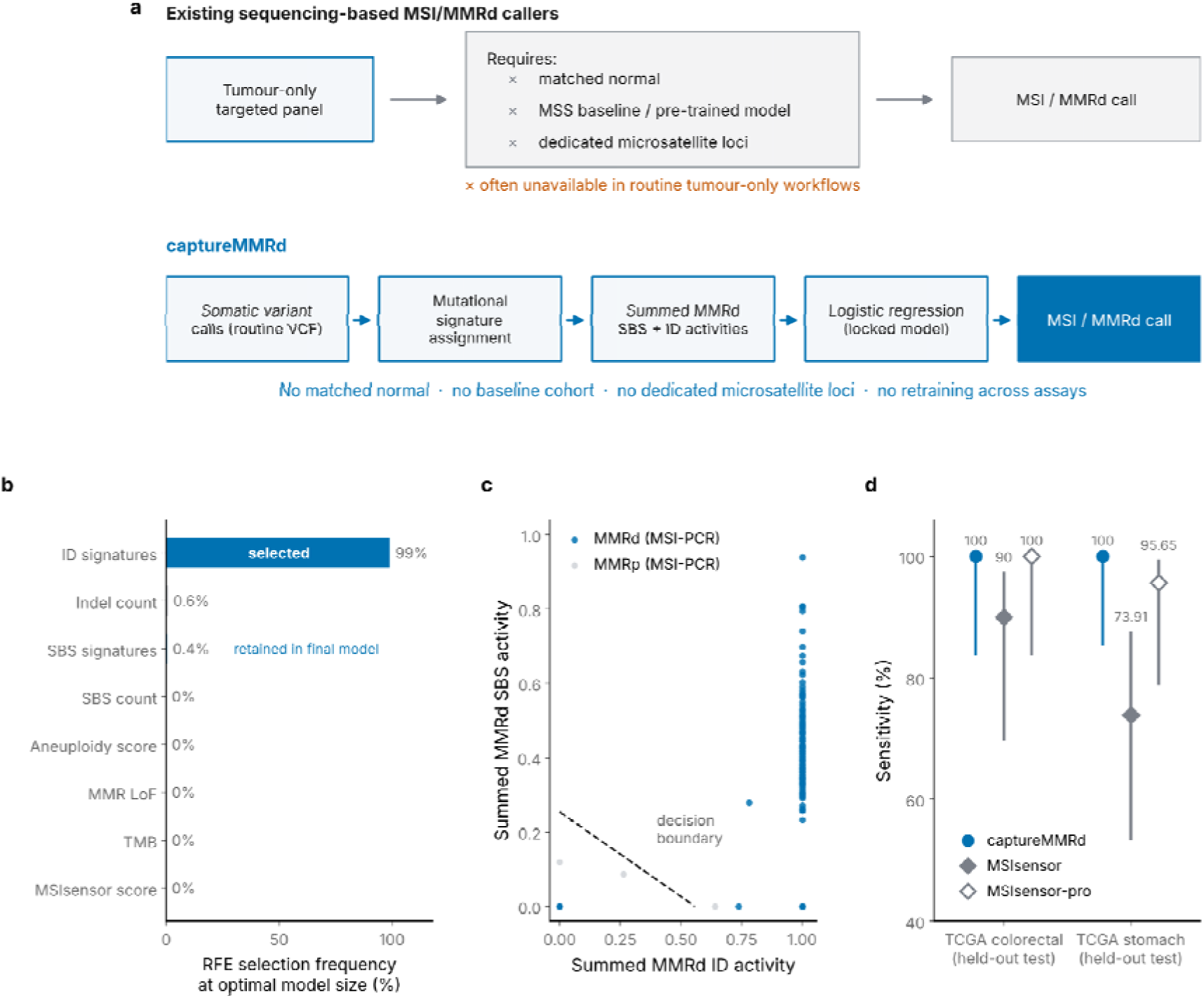
CaptureMMRd workflow, feature selection and model training and testing. a) CaptureMMRd workflow compared to traditional MSI/MMRd callers. b) Recursive feature elimination selection frequency c) CaptureMMRd decision threshold on MMRd SBS and MMRd ID signatures activities. d) CaptureMMRd sensitivity in the TCGA held-out test sets compared with MSIsensor and MSIsensor-spro. Error bars, 95% Wilson score confidence intervals.

## Results

### Mutational signatures are sufficient to diagnose MMRd from targeted sequencing

To identify the minimal genomic readout required for tumour-only MMRd diagnosis, we assessed a broad set of candidate NGS-derived features capturing complementary consequences of MMR loss: MSI scores, TMB, pathogenic alterations in the canonical MMR genes MLH1, PMS2, MSH2 and MSH6, aneuploidy, and SBS, DBS and ID mutational signatures associated with MMRd. For model development, we used the TCGA colorectal and stomach adenocarcinoma tumour–normal WES cohorts (**Supplementary Table 1**), with clinical MSI-PCR labels as the reference standard, as MSI-PCR is principally validated in gastrointestinal tumours^48^. After excluding cases with indeterminate reference results and ultramutated tumours carrying POLE/POLD1 alterations (Methods), cohorts were split at the patient level into training (70%) and held-out test (30%) sets, with all preprocessing and feature selection restricted to the training set. MMRd-associated SBS (SBS6, SBS14, SBS15, SBS20, SBS21, SBS26 and SBS44) and ID (ID1, ID2 and ID7) activities were summed into two aggregate features, with activity set to zero below an assay-specific minimum mutation count (563 SBS and 19 indels for WES) derived by exhaustive threshold optimization in the training set (Methods; Supplementary Tables 2 and 3).

Recursive feature elimination with cross-validation identified a model using ID signatures alone as optimal (Methods), with ID activity the selected feature in 99.0% of 500 training-set resamples (**Figure 1b**). Given that SBS and ID signatures are computed concomitantly at no additional cost, and that adding the SBS signature activity left the calls unchanged in 477 of those 500 resamples without reducing sensitivity or specificity (**Figure 1b**), we retained both signature classes, and trained a logistic regression classifier on the two aggregate signature activities (ID1+ID2+ID7 and SBS6+SBS14+SBS15+SBS20+SBS21+SBS44), a model we named captureMMRd. In training data, captureMMRd achieved a sensitivity of 95.56% and specificity of 99.36% in colorectal cancer, and 100% for both in stomach cancer (**Figure 1c**). Sensitivity and specificity values throughout the study are displayed along Wilson binomial intervals, reflecting cohort size for the fixed classifier with no model retraining.

In the held-out TCGA test sets, captureMMRd classified every sample correctly in both cohorts (sensitivity = 100%, 95% CI 83.89–100.00 in colorectal and 85.69–100.00 in stomach; specificity = 100%, 95% CI 97.19–100.00 in colorectal and 96.37–100.00 in stomach; **Figure 1d**). captureMMRd exceeded the sensitivity of MSIsensor (colorectal: 90.00%; stomach: 73.91%)^25^, and matched MSIsensor-pro in colorectal and exceeding it in stomach (colorectal: 100%; stomach: 95.65%)^28^ (**Figure 1d**). The final model was deliberately kept simple: two interpretable features and a linear classifier that can be locked, audited and deployed on any assay without retraining.

### A single locked model generalises across cohorts and assays without retraining

To determine whether captureMMRd generalises beyond its training domain, we applied the locked model, without retraining or threshold adjustment, to a series of increasingly distant validation settings: a tumour type unseen during training, independent non-TCGA cohorts, lower-depth WGS, and clinically implemented targeted panels (**Figure 2**).

**Figure 2:**
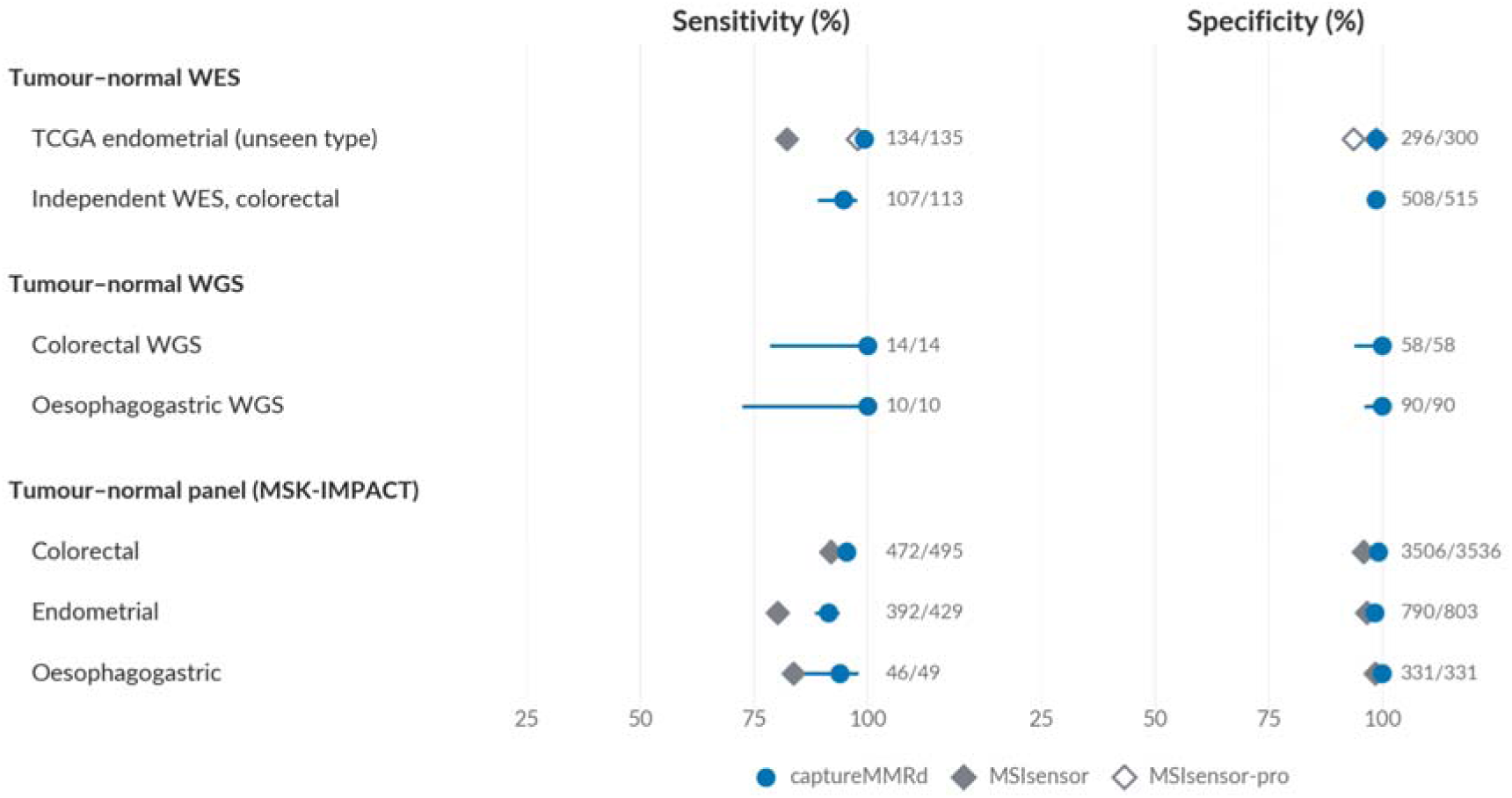
A single locked model generalises across cohorts and assays without retraining. Sensitivity and specificity of captureMMRd applied without retraining or threshold adjustment to 7 tumour-normal validation cohorts: TCGA uterine corpus endometrial carcinoma; an independent WES colorectal cohort; two WGS cohorts; and MSK-IMPACT targeted sequencing. MSIsensor and MSIsensor-pro are shown for comparison where available. Reference standard (MSI-PCR or MMR IHC) per cohort is given in Supplementary Table 1. Error bars, 95% Wilson confidence intervals.

CaptureMMRd achieved a sensitivity of 99.26% (134/135; 95% CI 95.92–99.87) and specificity of 98.67% (296/300; 95% CI 96.62–99.48) in TCGA uterine corpus endometrial carcinoma, a tumour type unseen during training (**Figure 2**), exceeding the sensitivity obtained with MSIsensor (82.22%) and MSIsensor-pro (97.78%). Subsets of non-colorectal MMRd tumours, including endometrial carcinoma, show fewer or smaller detectable microsatellite shifts than classical MSI-H colorectal cancers^49^. Since most traditional MSI assays and thresholds were optimised in colorectal cancer, this can lead to reduced sensitivity or discordant MMR results in endometrial cancer^13,50^. Reading mutational signatures instead of microsatellite loci appears to overcome this limitation.

To test generalisation beyond TCGA, we first evaluated captureMMRd in an independent tumour–normal WES validation cohort of 628 colorectal cancers, comprising 104 fresh-frozen^51^ and 524 FFPE^52^ samples with MSI-PCR status as the orthogonal reference standard. captureMMRd correctly identified 107/113 MSI-H tumours and 508/515 MSS tumours (sensitivity = 94.69%, 95% CI 88.90–97.54%; specificity = 98.64%, 95% CI 97.22– 99.34%; **Figure 2**). We next evaluated captureMMRd in tumour–normal WGS data. Across two independent cohorts, comprising 72 colorecta^58^ and 100 oesophagogastric^59^ tumour– normal pairs, the locked model showed complete concordance with the reference assays (24/24 MMRd; 148/148 MMRp; sensitivity = 100%, 95% CI 86.20–100.00; specificity = 100%, 95% CI 97.47–100.00; **Figure 2**). Finally we assessed captureMMRd in MSK-IMPACT, a clinically implemented tumour–normal targeted gene panel deployed across 341-, 410-, 468- and 505-gene versions. We aggregated a combined set of 5,643 colorectal, endometrial and oesophagogastric samples sequenced with MSK-IMPACT from eight different cBioPortal^53–56^ cohorts^30,56–62^ with available patient-level MMR IHC status. Without retraining or threshold adjustment, captureMMRd achieved sensitivities of 95.35% (472/495), 91.38% (392/429) and 93.88% (46/49) and specificities of 99.15% (3506/3536), 98.38% (790/803) and 100.00% (331/331) across these tumour types, outperforming MSIsensor in the same MSK-IMPACT datasets (**Figure 2**)

Taken together, our single locked model achieved a pooled sensitivity of 94.38% (1175/1245; 95% CI 92.96–95.53) and specificity of 99.04% (5579/5633; 95% CI 98.75–99.26) across 6,878 samples from 7 tumour–normal validation cohort–assay groups spanning WES, WGS and targeted panel sequencing (**Figure 2**). Having established that captureMMRd generalises across assay classes when a matched normal is available, we next asked whether it maintains this performance in the setting that motivated its development: tumour-only sequencing.

### Tumour-only panels and liquid biopsy support accurate MMRd diagnosis

Most routine clinical NGS is performed using tumour-only capture sequencing^44–47^, the setting in which existing sequencing-based MSI tools require panel-specific baselines, pre- trained models or dedicated microsatellite content. Tumour-only panels also vary widely in design, from standardised institutional assays to locally customised panels. We therefore evaluated captureMMRd in two independent tumour-only assays, an institutional panel deployed in three versions, a custom small panel, and in a plasma cfDNA assay.

In 453 colorectal adenocarcinomas sequenced with DFCI-OncoPanel^63^, a tumour-only institutional assay deployed in 275-, 300- and 447-gene versions, captureMMRd achieved a sensitivity of 92.86% (39/42; 95% CI 80.99–97.54) and specificity of 98.54% (405/411; 95% CI 96.85–99.33) against the MMR IHC reference standard (**Figure 3a**).

**Figure 3:**
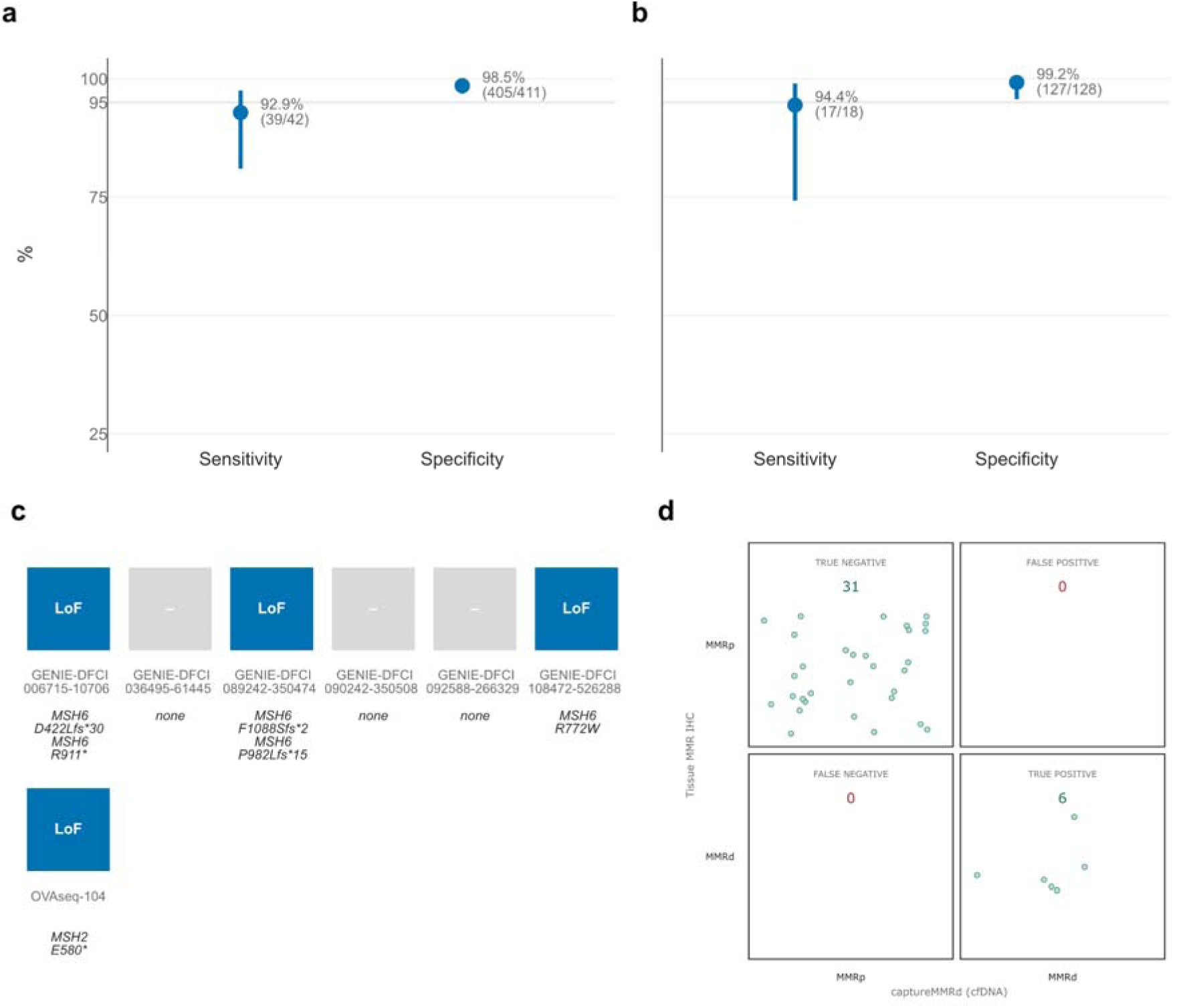
Tumour-only panels and liquid biopsy support accurate MMRd diagnosis. a) Performance in colorectal adenocarcinoma samples sequenced with DFCI-OncoPanel against MMR IHC. b) Performance in ovarian clear-cell and endometrioid carcinomas samples sequenced with the OVAseq panel against MMR IHC and MSI-PCR. c) Loss-of-function alterations in MMR genes among samples with discordant diagnosis between captureMMRd and the gold standard. d) Concordance with tissue MMR IHC in plasma cfDNA samples sequenced with MSK-ACCESS. Error bars, 95% Wilson confidence intervals.

To extend evaluation beyond the tumour types most commonly used for MMRd benchmarking, we profiled an institutional cohort of 146 ovarian clear-cell and endometrioid carcinomas–histotypes in which the clinical relevance of MMRd detection is increasingly recognised^64^–using a custom small targeted panel with MMR IHC and MSI-PCR as orthogonal reference assays. captureMMRd maintained strong performance despite the reduced panel size (∼0.5Mb), achieving a sensitivity of 94.44% (17/18; 95% CI 74.24–99.01) and specificity of 99.22% (127/128; 95% CI 95.71–99.86; **Figure 3b**).

Samples called MMRd by captureMMRd but not by the reference assays were enriched for loss-of-function alterations in MMR genes (**Figure 3c**). In the DFCI-OncoPanel cohort, three out of six such discordant samples harboured loss-of-function alterations in *MSH6*; in the ovarian cohort, the single discordant sample harboured loss-of-function alterations in *MSH2* (**Supplementary Table 4**). These observations suggest that a subset of discordant predictions reflect biologically plausible MMRd events missed by IHC or PCR, not false-positive calls, and support careful review of discordant cases before dismissal.

Finally, we tested captureMMRd in plasma cfDNA, a setting of increasing importance for patients with advanced or metastatic disease, where tissue availability may be limited, repeated biopsies are often impractical, and few approaches exist for MMRd detection. In 37 endometrial, colorectal and oesophagogastric adenocarcinomas with detectable ctDNA sequenced using MSK-ACCESS, a hybrid-capture cfDNA assay^65^ (Methods), captureMMRd was fully concordant with tissue-based MMR IHC (6/6 MMRd; 31/31 MMRp; **Figure 3d**), extending MMRd detection to liquid biopsy workflows.

In contrast to the discordant MMRd calls, the four samples called MMRp by captureMMRd despite an MMRd reference diagnosis (three in DFCI-OncoPanel, one in OVAseq, none in MSK-ACCESS) carried sparse mutation catalogues within the panel footprint: 0–10 somatic indels (median 5.5), compared with a median of 14 among correctly classified MMRd samples (two-sided exact Wilcoxon rank-sum test, p = 0.016; **Supplementary Table 8**). Two sat marginally below the decision threshold, with summed MMR indel activity of 0.50 against an effective threshold of 0.56. The OVAseq false negative, MSH6-deficient by IHC, carried no somatic indels and four SBS within the panel, and was also classified as microsatellite stable by MSIsensor2 (score 9.1), indicating an absence of instability signal in the sequencing data itself. Two of the four false negatives were MSH6-deficient, a genotype associated with an attenuated instability phenotype^66^. The sensitivity limit of captureMMRd is therefore set by the number of informative somatic mutations captured by the panel, a constraint shared by sequencing-based approaches at low mutation burden.

Across all validation settings combined, a single locked model achieved a pooled sensitivity of 94.36% (1237/1311; 95% CI 92.97–95.48) and specificity of 99.02% (6142/6203; 95% CI 98.74–99.23) across 7,514 samples from 16 independent cohorts, four assay classes and five tumour types (**Figures 2 and 3**). captureMMRd thus extends accurate MMRd diagnosis to the tumour-only panels and liquid biopsy assays used in routine precision oncology, using only the somatic variant calls these assays already capture.

### captureMMRd matches tumour-only MSI callers without requiring microsatellite loci

Existing approaches to tumour-only MSI/MMRd detection fall into two classes: microsatellite-locus readers, which depend on the panel’s microsatellite content and platform-specific calibration, and single-feature mutation heuristics, exemplified by homopolymer indel counts calibrated against IHC in colorectal cancer^36^. These classes differ in what they require as input. Microsatellite-locus callers operate on aligned sequencing reads, available in this study only for the OVAseq cohort, whereas the indel-rate rule requires only the somatic variant calls available for every cohort. We therefore evaluated the indel-rate rule across all cohorts and performed a head-to-head comparison of all methods in OVAseq to test each class’s dependence on assay design under controlled conditions.

Applied across cohorts using each panel’s exonic footprint, the indel-rate rule showed similar sensitivity (100.0%, 94.4% and 100.0%; **Figure 4a**), but poorer specificity compared to captureMMRd (89.54%, 96.88% and 96.77%; **Figure 4a**). The sbs-rate rule instead showed much lower sensitivity compared to captureMMRd (73.8% and 33.3%, sbs-rate rule was not applied to MSK-ACCESS as no sample in the cohort had enough sbs; **Figure 4a**), but equivalent specificity in the cohorts tested (99.0% and 99.2%, sbs-rate rule was not applied to MSK-ACCESS as no sample in the cohort had enough sbs; **Figure 4a**).

**Figure 4:**
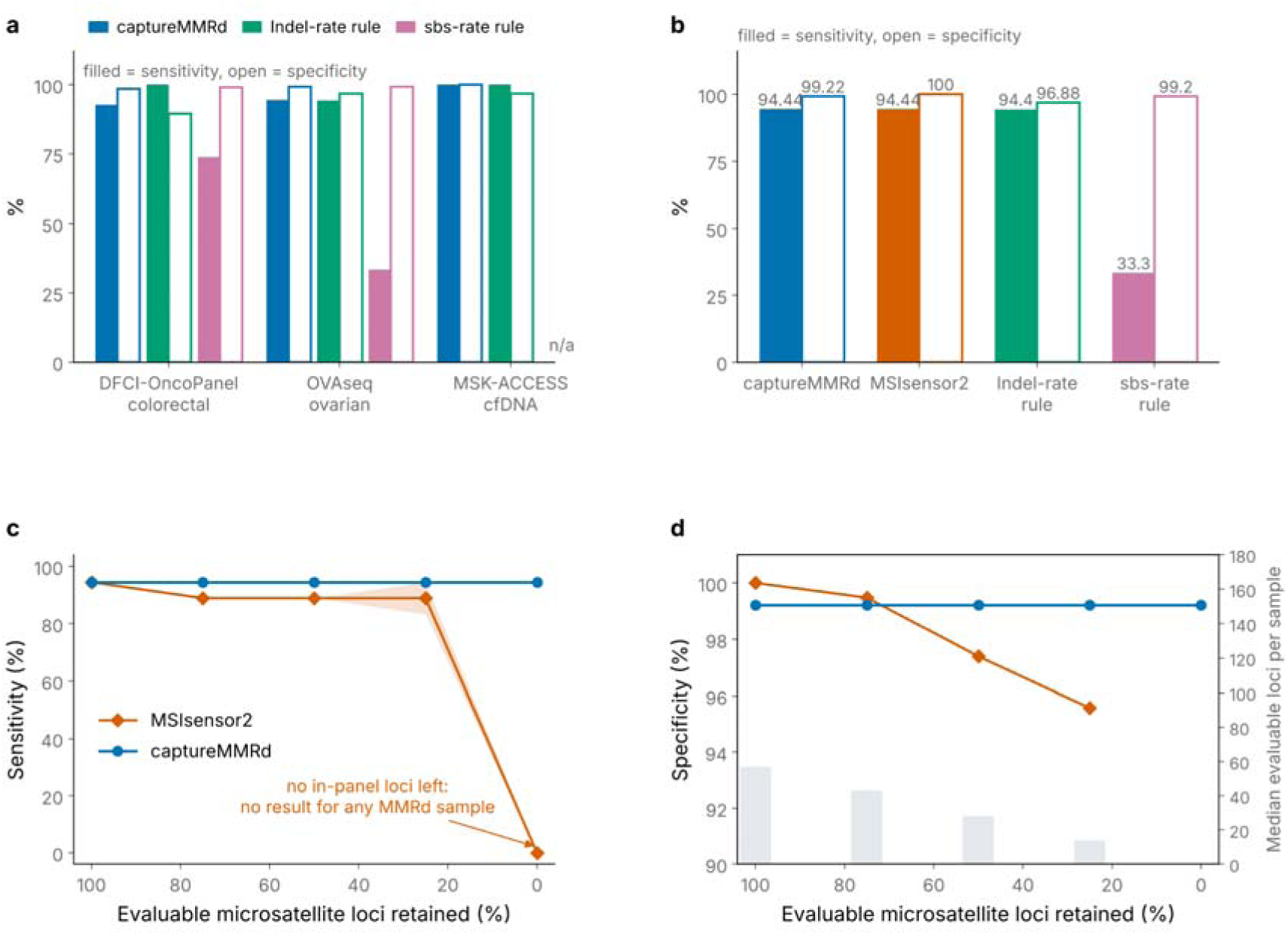
captureMMRd matches tumour-only MSI callers without requiring microsatellite loci. a) Sensitivity and specificity of the indel-rate, sbs-rate rule and captureMMRd applied across tumour-only and liquid biopsy cohorts. b) Head-to-head comparison in the OVAseq cohort of captureMMRd, MSIsensor2 and the indel-rate rule on identical samples. c) Sensitivity of MSIsensor2 and captureMMRd in the OVAseq cohort (n = 146; 18 MMRd) under in silico removal of microsatellite-bearing targets, retaining 100%, 75%, 50%, 25% and 0% of evaluable microsatellite loci. Points, mean across three random removal orderings; shading, range across orderings. At 0% retained loci, MSIsensor2 returns no result for any MMRd sample. d) Specificity under the same reductions; bars, median evaluable microsatellite loci per sample.

In OVAseq, the cohort with raw sequencing data, we compared captureMMRd directly against MSIsensor2, a pre-trained microsatellite-locus caller^29^ on identical samples. captureMMRd achieved a sensitivity of 94.44% (17/18) at a specificity of 99.22% (127/128), compared with 94.44% (17/18) and 100% (128/128) for MSIsensor2, the two methods separated by a single discordant MMRp sample (**Figure 4b**). As before, the indel-rate and sbs-rate rules fell below both methods (**Figure 4b**).

To determine whether this equivalence depends on the panel’s microsatellite content, we progressively removed microsatellite-bearing targets from the OVAseq designs *in silico* and re-ran both methods on identical reductions. The OVAseq panels were designed to enrich microsatellite content, through dedicated microsatellite regions and a subset of exonic targets; in total, 63–65 targets per panel version, 2.3–2.5% of the panel footprint, contained every locus evaluable by MSIsensor2 (23–66 loci per sample passing its coverage filter). We removed these targets in ranked order to retain 75%, 50%, 25% and 0% of evaluable loci, across three random removal orderings per level (Methods), excluding the variants they contained from signature estimation.

MSIsensor2 performance degraded with panel design: mean sensitivity fell from 94.4% (17/18) on the full panel to 88.9% across the depletion levels (range 83.3–94.4% at 25% retained loci), specificity fell to a mean of 95.6%, and with every microsatellite-bearing target removed it returned no result for any MMRd sample (**Figure 4c,d**). captureMMRd diagnoses were unchanged throughout: all 146 samples received the same call as on the full panel at every depletion level, including complete removal, at a cost of less than 2.5% of the panel footprint (**Figure 4c,d**). The two methods therefore fail in different ways: a microsatellite-locus caller fails through panel design, whereas captureMMRd fails only when too few mutations remain to estimate signature activity, a condition we characterise below.

### captureMMRd is robust to preservation method and reduced tumour purity

Clinical deployment requires stability against the variables that differ most across laboratories. Sample preservation is one of these variables that could impact the quality of samples the most. Specifically, formalin fixation introduces characteristic artefactual substitutions that could in principle contaminate signature estimates. The MMRd-associated signature set does not include FFPE-artefact processes, and performance did not statistically differ between fresh-frozen (sensitivity=100% and specificity=100%) and FFPE (sensitivity=93.33% and specificity=98.39%) samples in the independent WES cohort (Fisher’s exact, p = 0.344 for sensitivity and p = 0.603 for specificity) (**Figure 5**).

**Figure 5:**
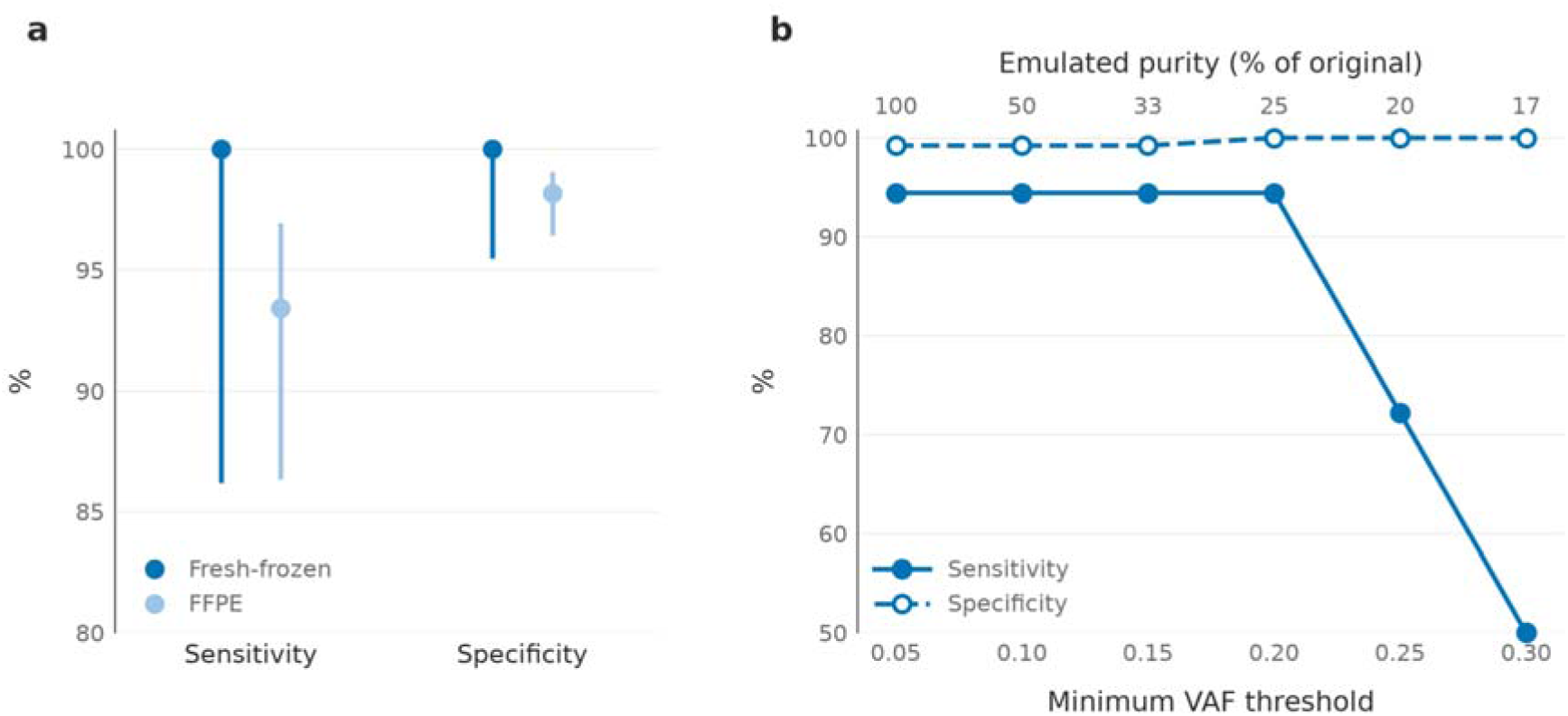
captureMMRd is robust to preservation method and reduced tumour purity. a) Performance by sample preservation type (fresh-frozen versus FFPE) in the tumour-normal WES cohorts; error bars, 95% confidence intervals. b) Sensitivity and specificity in the OVAseq cohort (n = 146; 18 MMRd) as variants are re-filtered at rising minimum VAF thresholds, from 0.05 (the standard filter) to 0.30; top axis, equivalent tumour purity relative to each sample’s original, assuming a 5% VAF detection limit.

Tumour purity varies widely in clinical specimens and lowers the variant allele fractions (VAFs) at which somatic mutations are observed. To emulate falling purity, we re-filtered the OVAseq cohort at minimum VAF thresholds from 0.05 (baseline) to 0.30 and repeated signature assignment and classification (Methods). Diagnoses were unchanged at a threshold of 0.10 (146/146 concordant with baseline), and the only changed calls at thresholds of 0.15 and 0.20 moved toward the reference standard, with the single baseline false positive reverting to MMRp (94.44% sensitivity at 100% specificity at 0.20). From a threshold of 0.25, equivalent to approximately 20% tumour purity at a 5% detection limit, sensitivity declined (14/18, and 11/18 at 0.30) as indel counts fell below the minimum mutation threshold. As with panel composition, reduced purity degrades captureMMRd only through the predictable minimum-mutation failure mode.

### Minimal panel requirements for MMRd diagnosis

The robustness analyses identified a single predictable failure mode: too few mutations to estimate signature activity. Because captureMMRd relies on the SBS and indels captured within a panel’s footprint, small panels may yield mutation counts below the reliable range, not through any property of the tumour but through assay design. We therefore characterised the minimum input required for a reliable diagnosis, and translated it into a prospective eligibility rule for any panel.

Using the TCGA training cohort, we fixed the number of SBS and ID mutations available per sample and evaluated prediction stability at each count. Three indels were sufficient to reach a 90% agreement with the full indel set in MMRp and MMRd samples (**Figure 6a**). For SBS signatures, because they were already highly specific, we just focused on MMRd samples. Here, 44 mutations were required to reach the 90% accuracy agreement (**Figure 6b**). These minima reflect the information content of each mutation class: indels at short repeats are the most direct consequence of MMR loss, so each observed indel carries substantially more diagnostic weight than a substitution.

**Figure 6:**
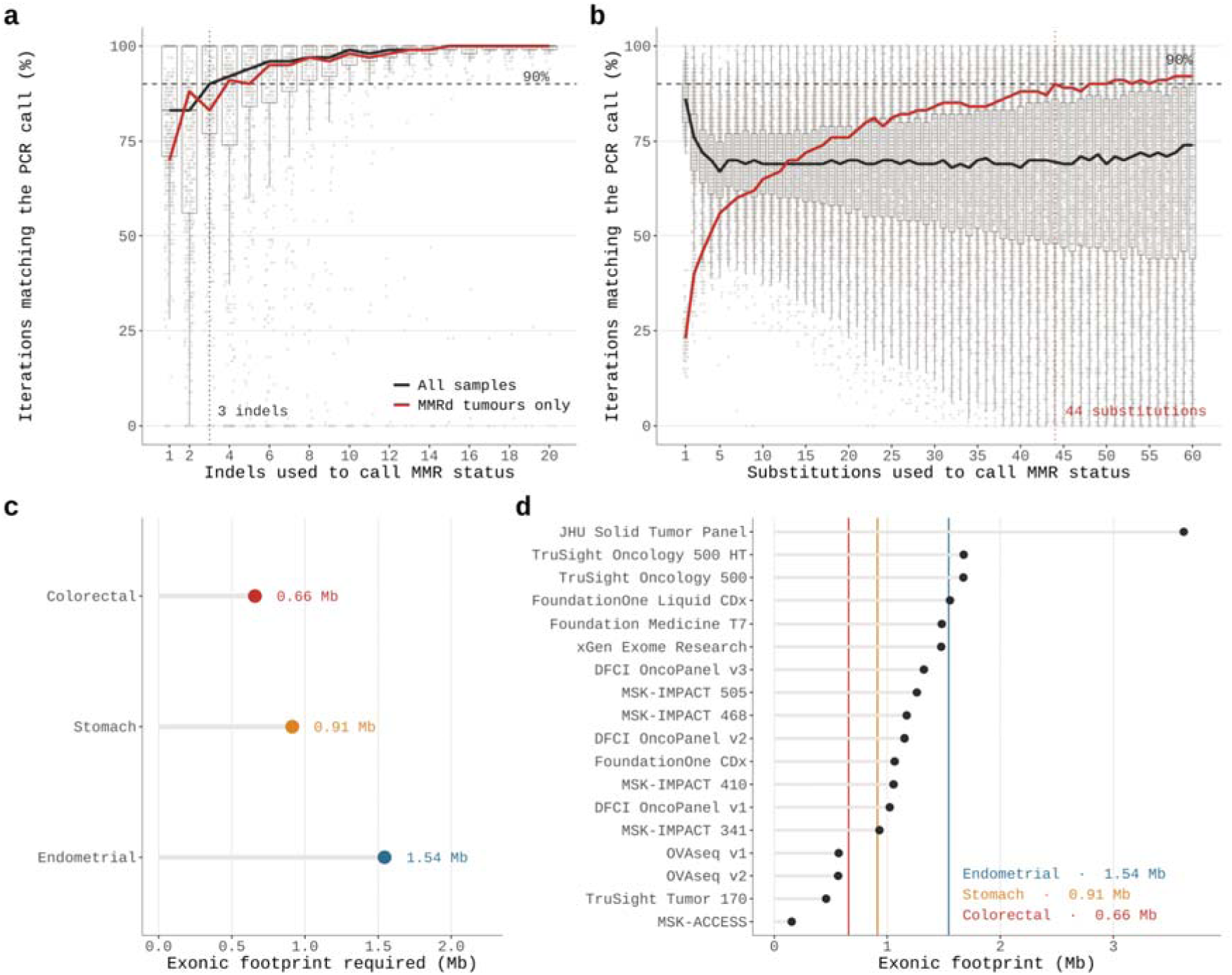
Minimal panel requirements for MMRd diagnosis. a) Accuracy of captureMMRd as a function of the number of indels used per sample in the TCGA training cohort. b) Accuracy of captureMMRd as a function of the number of sbs used per sample in the TCGA training cohort. c) Minimum exonic panel footprint (Mb) required for reliable diagnosis in colorectal, stomach and endometrial cancer. d) Exonic footprints of widely known targeted panels in current clinical use relative to these cancer types minima (Supplementary Table 5).

We next translated these minima into panel size. Combining the required mutation counts with the per-megabase ID rates (because indels alone carry most of the MMRd signal) of MMRd tumours in each cancer type, a panel footprint of 0.66Mb is sufficient for reliable MMRd diagnosis in colorectal, 0.91Mb in stomach cancer, and 1.54Mb in endometrial cancer (**Figure 6c**). The difference across tumour types reflects their distinct baseline mutation rates, and the estimates are deliberately conservative, derived from the lower range of mutation burdens observed among MMRd tumours.

To assess what these requirements mean in practice, we surveyed the exonic footprints of targeted panels which are widely used in the clinical setting, spanning commercial assays, institutional panels and the designs evaluated in this study (**Figure 6d; Supplementary Table 5**). Most panels exceed the minimum footprint for colorectal and stomach cancer, while endometrial cancer might require more regions covered. Overall, this indicates that most clinically deployed assays could support captureMMRd without modification for colorectal and stomach cancers. Panels below the threshold are identifiable in advance from their design files alone, requiring no sequencing to establish eligibility.

## Discussion

In this study, we demonstrate that mismatch repair deficiency can be diagnosed from the somatic variant calls produced by routine tumour-only sequencing, using mutational signatures alone. A single locked model, trained once on tumour-normal WES, maintained a pooled sensitivity of 94.36% and specificity of 99.02% across 7,514 samples from 16 independent cohorts without retraining, recalibration or access to matched normal DNA. Previous sequencing-based approaches have anchored MMRd detection to the assay: microsatellite-locus callers require matched normal sequencing^25,26^, panel-specific baselines^27,28^, platform-specific pre-training^29,30^ or dedicated microsatellite content^19,31–33^, and the only mutation-based alternative in clinical use is a single-feature indel-rate rule calibrated within one institutional colorectal workflow^36^. captureMMRd generalises the biology these approaches sample into a panel-agnostic classifier: the mutational consequences of MMR loss are read directly from the variant calls, so any panel that captures enough somatic mutations can support a diagnosis.

Framing detection around signatures changes where the method can fail. In the one cohort with raw sequencing data, captureMMRd and MSIsensor2 were separated by a single sample, yet the two methods degraded differently when microsatellite-bearing targets were removed in silico: MSIsensor2 lost sensitivity progressively and returned no result once its loci were gone, while captureMMRd calls were unchanged at every depletion level, at a cost of less than 2.5% of the panel footprint. A locus caller fails through panel design, sample by sample, as informative content shrinks. captureMMRd fails only when too few mutations remain to estimate signature activity, a condition that is explicit in the model, predictable from a panel’s design file before any sequencing is performed, and reported as an eligibility criterion instead of a silent miscall. Calls were similarly stable to variant-filtering stringency, which we used to emulate reduced tumour purity, and to preservation method. Samples called MMRd by captureMMRd but not by the reference assays were enriched for loss-of-function alterations in MMR genes, consistent with the known failure modes of IHC and MSI-PCR^11–13^; signature-based readouts may therefore recover a subset of MMRd tumours that current standards miss, although orthogonal confirmation would be required before such calls direct treatment.

We see captureMMRd as a complement to established testing, not a replacement for it. IHC and MSI-PCR remain the clinical standard where they are available and validated^8–10^. The setting captureMMRd addresses is the one existing methods leave uncovered: laboratories running tumour-only panels, particularly smaller or custom designs, that lack the matched normals, baseline cohorts and microsatellite content existing sequencing-based methods assume. For these laboratories, MMRd status becomes computable from data they already generate, at no additional assay cost, with eligibility determinable in advance from the panel design alone. The same property permits retrospective re-analysis: archived variant calls from historical cohorts can be screened for MMRd without returning to tissue, with implications for Lynch syndrome ascertainment^4,5^ and for identifying patients who may benefit from immune checkpoint blockade^1–3^. The full concordance observed in plasma cfDNA further suggests a route to MMRd assessment where tissue is unavailable, although this requires validation beyond the 37 samples tested here.

Our study has limitations. We treated IHC and MSI-PCR as ground truth, so where the reference errors, our reported sensitivity and specificity are conservative; we could not resolve the discordant calls with an independent assay. Training used colorectal and stomach adenocarcinoma with MSI-PCR labels, which are principally validated in gastrointestinal tumours^48^, and validation, although pan-cancer in aggregate, is dominated by the tumour types common in clinical sequencing cohorts; performance in rarer types, and in endometrial cancer on small panels, where the minimum footprint of 1.54 Mb exceeds several deployed designs, requires further evaluation. Raw sequencing data were available only for the OVAseq cohort, so the head-to-head comparison and the robustness analyses are confined to one assay, and the depletion and purity analyses were performed in silico on panel designs; prospective sequencing on panels without microsatellite content has not been performed. Finally, signature fitting near the minimum mutation counts is intrinsically unstable^67–69^. We handle this with explicit per-assay minima, assigning zero activity below them, which renders a subset of samples on very small panels ineligible instead of miscalled, and this indeterminate fraction is a cost of the approach that laboratories should report alongside performance.

Further work will be necessary to establish performance prospectively across laboratories and bioinformatic pipelines, to extend validation to the tumour types and panel designs underrepresented here, and to define how signature-based calls should be combined with IHC and MSI-PCR in diagnostic algorithms. By operating on the output every sequencing workflow already generates, captureMMRd extends clinically actionable MMRd detection to assays that were never designed to measure it. The broader principle is that DNA repair deficiencies leave mutational scars that routine panel data already record; reading them directly may bring biomarker-directed therapy to every setting where targeted sequencing is performed.

## Methods

### Cohorts

We analysed samples from 18 independent cohorts comprising different sources, specimen types, sequencing assays, sample design and gold standard diagnosis.

Tumour-normal fresh frozen WES data was also retrieved from cBioPortal for three cohorts from The Cancer Genome Atlas (TCGA): Colorectal Adenocarcinoma (*coadread_tcga_pan_can_atlas_2018*), Stomach Adenocarcinoma (*stad_tcga_pan_can_atlas_2018*), and Uterine Corpus Endometrial Carcinoma (*ucec_tcga_pan_can_atlas_2018*). MMR-D status in these cohorts was defined using MSI PCR. Additionally, three more tumour-normal WES non-TCGA cohorts containing fresh frozen or FFPE samples were included in which either MSI PCR alone or together with MMR IHC was used as MMR status gold standard diagnosis ^51,52,70^.

Tumour-normal fresh frozen WGS data was extracted for Colorectal ^71^ and Stomach Adenocarcinoma ^72^ from cBioPortal ^53^ ^54,55^. MRR-D diagnosis regarding Colorectal Adenocarcinoma was assessed using either MMR IHC or MSI PCR, while for Stomach Adenocarcinoma only MSI PCR was used.

Pancancer targeted tumour-normal sequencing data were obtained from MSK-IMPACT MiMSI cohort (*pancan_mimsi_msk_2024*), also available through cBioPortal ^53^. MMR-D labels for these cohorts were defined based on available IHC or PCR results. Moreover, we also obtained MSK-IMPACT Colorectal Cancer data from the MSK-CHORD cohort (*msk_chord_2024*), from the Metastatic Colorectal Cancer cohort (crc_msk_2017) and from the Nigerian Colorectal Cancer cohort (*crc_nigerian_2020*). IHC and PCR were used as gold standard diagnosis respectively for these cohorts. Endometrial cancer data sequenced with MSK-IMPACT was also retrieved from the MSK-UCEC cohort (*msk_ucec_2018*), from the Endometrial Carcinoma MSI cohort (ucec_ccr_msk_2022) and from the Endometrial and Ovarian Cancer cohort (ucec_msk_2024). MMR IHC was used as the reference diagnosis for these samples.

Colorectal cancer tumour-only data was retrieved from samples sequenced with the DFCI-OncoPanel from the GENIE BioPharma Collaborative (BPC) colorectal cancer (CRC v2.0-public) cohort (*crc_public_genie_bpc*)^55^ from the publicly available AACR Project GENIE database^56^ as well as from cBioPortal ^73^. Additionally, colorectal adenocarcinoma tumour-only data sequenced with DFCI-OncoPanel, was also retrieved from cBioportal (crc_orion_2024). Somatic mutation data were used to derive mutational signatures, while mismatch repair immunohistochemistry (MMR IHC) and microsatellite instability PCR (MSI-PCR) results were used as reference standards for MMR deficiency classification.

Analyses of publicly available data from TCGA, AACR Project GENIE, MSK and the other published cohorts were performed under the terms of access of each resource and required no additional approval.

For circulating tumour DNA (ctDNA), we used the MSK ctDNA sequencing cohort (*msk_ctdna_vte_2024*) dataset that uses the MSK-ACCESS platform, selecting only those samples with matched tissue MMR IHC results reported in either MSK-CHORD (msk_chord_2024) or MiMSI cohort. Because total cfDNA input for MSK-ACCESS library preparation was not available, we used plasma cfDNA concentration as a surrogate for sample adequacy. MSK-ACCESS was validated using 20 ng of plasma cfDNA whenever possible, with analyses attempted down to 3 ng. We therefore applied a 2.5 ng/mL cutoff to select samples likely to provide near-optimal cfDNA input, assuming plasma volumes similar to those commonly obtained in clinical liquid biopsy workflows, where 8–10 mL of plasma would yield approximately 20–25 ng cfDNA at this concentration. Thus, only samples with cfDNA concentrations ≥2.5 ng/mL were included in this analysis. Because samples below this threshold were not analysed, we cannot assess the performance of the method in lower-yield cfDNA samples. Additionally, we only considered samples with max VAF>0.005, as this was the VAF threshold at which MSK-ACCESS is reported to display 92% sensitivity and a 99% specificity for de novo SNVs and indels calling ^65^

Finally, we evaluated an institutional cohort of 146 ovarian tumour samples from a multicentric study, sequenced with a custom tumour-only panel. MMR-D status was determined by MMR IHC or MSI-PCR. Samples from this cohort were provided by the IdiPAZ Biobank (PT23-00028), integrated into the ISCIII Biobanks and Biomodels Platform; the Biobank of the Andalusian Public Health System (SSPA), which coordinated the collection, processing, management and distribution of the biological samples in accordance with its established standard procedures; and the BioBank of the Fundación MD Anderson Cancer Center España-Hospiten, integrated into the Spanish National Biobanks Network (B.0000745). All samples were processed following the standard operating procedures established by the respective biobanks, with the appropriate approval of their Ethics and Scientific Committees. This study was conducted in accordance with ethical standards of the Helsinki Declaration of the World Medical Association and ethical approval was obtained from the ISCIII Research Ethics Committee (CEI PI 96_2019-v2) and conducted in accordance with the Declaration of Helsinki.

Of note, we removed all samples with indeterminate gold standard diagnosis for training, testing and validation of the model. Additionally, we followed the clinical guidelines for commercial targeted panels used in this study and restricted the analysis of MSK-IMPACT samples to: tumour purity > 10%, sample coverage > 200x, and matched somatic status. Whereas for DFCI-OncoPanel we just evaluated samples with: tumour purity > 20%. No purity or coverage filter was applied to WES, WGS, OVAseq or the cfDNA cohort. This included samples with single test indeterminate results and multiple but discordant gold standard diagnosis either being repeated discordant single IHC/PCR tests or both IHC-PCR discordancy. Ultramutated samples, either displaying TMB>100 or LoF mutations in POLE or POLD1, were also excluded for model training, testing and validation. These cancers have a clear path in the clinic, usually being referred to immunotherapy, independently of their MMR status. But because they share common features with MMRd cancers including them in the training might bias model performance. Additionally, patient-level deduplication is applied to the whole validation database after merging all cohorts data. Finally, MSI-L is folded into MSS before any modelling for any cohort reporting MSI-L in the MSI-PCR assay.

A summary of all cohorts is present at Supplementary Table 1.

### NGS data generation

#### Sequence panel design

We used a custom targeted sequencing panel comprising frequently altered cancer driver genes and 53 additional microsatellite regions. Two versions of the panel were used: version 1 targeted 145 genes (Supplementary Table 6), whereas version 2 targeted 155 genes (Supplementary Table 7). The total target regions covered approximately 0.57 Mb and 0.57 Mb (565,472 bp and 570,483 bp of target space), respectively. Capture probes were designed against the GRCh38 human reference genome.

#### Genomic DNA extraction and sample sequencing

For the institutional OVAseq cohort, genomic DNA was extracted from FFPE tumour samples using either the Maxwell RSC DNA FFPE Kit (Promega; catalog no. AS1450) or the truXTRAC FFPE Kit (Covaris; catalog no. 520136).

Library preparation differed between panel versions. Libraries for version 1 were prepared using the SeqCap EZ HyperCap Kit (Roche; catalog no. 08278512001), whereas libraries for version 2 were prepared using the KAPA HyperCap Kit (Roche; catalog no. 07962428001), in both cases according to the manufacturer’s instructions. Libraries generated using version 1 were sequenced on Illumina HiSeq 2500 or HiSeq 4000 instruments using 101-bp and 150-bp paired-end reads respectively. Libraries generated using version 2 were sequenced on an Illumina HiSeq 2000 instrument using 110-bp paired-end reads.

### Bioinformatic Analysis

For the OVAseq cohort, paired-end FASTQ files were processed using nf-core/sarek v3.8.1^74,75^, implemented in Nextflow^76^ within the nf-core framework^77^. Reads were aligned to the GRCh38 human reference genome using BWA-MEM2 v2.2.1^78^.

Sequencing quality-control metrics were calculated using Picard CollectHsMetrics v3.1.1. Samples were retained when at least 90% of targeted bases had a sequencing depth of ≥30× (PCT_TARGET_BASES_30X ≥ 0.90), median target coverage was ≥50× (MEDIAN_TARGET_COVERAGE ≥ 50), and the proportion of targets with zero coverage was ≤0.005 (ZERO_CVG_TARGETS_PCT ≤ 0.005). Patients with more than one sample were deduplicated selecting the first sample received from the ones that passed the QC. Overall, 146 OVAseq deduplicated samples passed these quality-control criteria and were evaluated.

#### Variant calling

For all publicly available cohorts, variant calls were obtained directly from their corresponding repositories. These variants were used with the annotations provided by each repository. For each cohort, information regarding the reference genome, variant-calling pipeline, and source repository is provided in (Supplementary Table 1).

For the OVAseq cohort, variants were called using nf-core/sarek pipeline v3.8.1^74,75^. This pipeline uses Mutect2 from GATK v4.6.1.0^79,80^, FreeBayes v1.3.10 ^81^, LoFreq v2.1.5 ^82^, and BCFtools v1.21^83^. Consensus variants were defined as variants detected by at least 2 callers. Variants were annotated with the Ensembl Variant Effect Predictor (VEP) ^84^.

#### Definition of predicted loss-of-function alterations

For publicly available cohorts, variants were classified as clinically relevant alterations if they were annotated as “Pathogenic” or “Likely pathogenic” in ClinVar^85^ or “Oncogenic” or “Likely oncogenic” in OncoKB ^86,87^.

For the OVAseq cohort, variants were filtered to identify predicted loss-of-function alterations. Variants were required to be rare in the general population (1000 Genomes^88^ allele frequency <1% and not common in ExAC^89^), have sufficient sequencing support (variant allele frequency >5%, ≥5 alternative-supporting reads, and total depth ≥10 reads), and affect a canonical transcript. SBS mutations were additionally restricted to variants called by at least 3 variant callers and to have a described reference SNP cluster ID whenever it was detected in more than 3 samples in our cohort. Predicted LoF alterations included frameshift, stop-gain, start-loss, and essential splice-site variants. Non-truncating exonic variants were also retained when classified as “Pathogenic” or “Likely pathogenic” in ClinVar.

#### Indel mutational signatures calling

For the COAD, STAD and UCEC TCGA cohorts we retained only variants that (1) passed filtering (FILTER ∈ {PASS, wga, native_wga_mix}) and (2) were annotated as canonical (CANONICAL = YES). For all the other publicly available cohorts, mutational matrices were computed using the raw mutations retrieved from each repository without any filtering. Regarding our institutional cohort, OVAseq, variants were included, expanding the criteria for the predicted LoF to also include synonymous mutations.

Mutational matrices were generated using SigProfilerMatrixGenerator v1.3.6 ^90^ and translated into SBS and ID mutational signatures using SigProfilerAssignment v1.1.3 ^67^ over the v3.5 of COSMIC signatures ^37^ ^91^.

#### Microsatellite instability computation

For the TCGA and MSK-IMPACT cohorts, MSIsensor scores were obtained from cBioPortal. For TCGA samples, MSIsensor-pro scores were additionally obtained from the supplementary data associated with ^28^. MSIsensor is called MSI-H at score ≥ 10, with scores between 3 and 10 treated as indeterminate and counted as a non-MSI-H cal and MSS at scores lower than 3l; MSIsensor-pro has no published cut-off, so the score maximising balanced accuracy at the training set was chosen, locked and applied unchanged.

For the OVAseq cohort, microsatellite instability was quantified from tumour-only sequencing data using MSIsensor2. As suggested by the authors, a score of 20% was used to classify samples as MSI-high when a categorical classification was required ^29^.

#### In silico microsatellite-target depletion

The OVAseq panel designs include targets selected for microsatellite content, in dedicated microsatellite regions and in a subset of exonic targets. For each panel version, every microsatellite locus evaluable by MSIsensor2 (≥20 tumour reads) was assigned to its covering target; 63–65 targets per version (2.3–2.5% of the panel footprint) contained all evaluable loci. Targets were removed in descending order of evaluable-locus count, with ties broken at random across three seeds, until 75%, 50%, 25% and 0% of in-panel evaluable loci were retained. Each resulting design was applied identically to both methods. For MSIsensor2, per-sample scores were recomputed from its per-site output restricted to retained loci (evaluable, ≥20 tumour reads; score, percentage of evaluable loci called somatic; MSI-H, score ≥20%); samples with no remaining evaluable locus were counted as returning no result. For captureMMRd, variants within removed targets were excluded, SBS96 and ID83 catalogues rebuilt, COSMIC v3.5 signatures refit with SigProfilerAssignment, and the locked model reapplied including the minimum mutation thresholds. Depletion performance is reported as concordance with each method’s full-panel calls within the same pipeline.

#### VAF-dilution purity emulation

To emulate reduced tumour purity, the filtered OVAseq variant set was re-thresholded at minimum VAFs of 0.05 (the baseline filter), 0.10, 0.15, 0.20, 0.25 and 0.30. At each threshold, SBS96 and ID83 catalogues were rebuilt from the retained variants, COSMIC v3.5 signatures refit with SigProfilerMatrixGenerator and SigProfilerAssignment, and the locked model reapplied including the minimum mutation thresholds. A threshold t with a baseline detection limit of 0.05 emulates a tumour purity of approximately 0.05/t. Performance is reported as call concordance with the baseline threshold.

#### Tumour mutational burden measurement

For the publicly available cohorts, tumour mutational burden (TMB) values were obtained from cBioPortal. We used the “TMB (nonsynonymous)” attribute for TCGA cohorts and the “Impact TMB Score” attribute for MSK-IMPACT cohorts. Values were analysed as provided by cBioPortal and expressed as mutations per megabase (mut/Mb).

For the OVAseq cohort, TMB was calculated as the number of qualifying nonsynonymous coding variants divided by the panel-specific exome-targeted territory, expressed in megabases. Variants were required to match the same filters as for the predicted LoF mutations. Because two panel versions were used, separate TMB denominators were calculated. For each version, the panel target intervals were intersected with the Genecode protein coding sequences (gencode.v49.basic.protein_coding.CDS.merged.bed). Overlapping intervals were merged to avoid counting individual bases more than once. The resulting exome-targeted territories were 0.53 Mb for panel version 1 and 0.55 Mb for panel version 2.

#### Aneuploidy scores

Aneuploidy scores for the TCGA cohorts were extracted from cBioPortal.

### Machine learning classifier

#### Data preprocessing

TCGA stomach adenocarcinoma (STAD) and colorectal adenocarcinoma cohorts (COADREAD) were used for model learning. MSI-PCR was used as the gold standard label for the model. Because MSI-PCR is principally validated for gastrointestinal tumours and is not the recommended reference assay for endometrial cancer ^48^, UCEC was used for model validation. Hence, STAD and colorectal cohorts were stratified by cohort and divided at the patient level into training (70%) and test (30%) sets using a stratified random split that preserved the proportion of MMRd and MMR-proficient samples. All preprocessing and feature-selection procedures were restricted to the training set.

We developed a binary classifier to predict mismatch repair deficiency (MMRd) from NGS-derived biomarkers. The candidate features were LoF mutations in the MMR genes, MSIsensor score, TMB, Aneuploidy Score, SBS Signatures (Sum of SBS6, SBS14, SBS15, SBS20, SBS21, SBS26 and SBS44), Indel Signatures (Sum of ID1, ID2 and ID7) associated with MMR-D and the per-sample indel and SBS catalogue sizes. LoF mutations in the MMR genes was a binary variable and was therefore not scaled. SBS and ID signatures were not scaled either as their values already ranged from 0 to 1. Continuous features such as MSIsensor score, TMB, sbs counts, indel counts and Aneuploidy Score were transformed using min–max normalization exclusively using the training data. This prevented information leakage and ensured that features contributed comparably to coefficient-based elimination. Samples with missing features or reference-standard data were excluded.

#### Minimum number of SBS and ID to call mutational signatures per technology

Because of the compositional nature of mutational signatures^37,92^, just a few random similar-context variants can lead to high signature activities, even though they might not be truly associated with any mutational process, and no established per-sample minimum mutation count exists for signature assignment^67–69^. Hence, we performed exhaustive threshold optimization in the training set over the number of SBS and ID mutations per-sample to find the minimum number of mutations to reliably call SBS and ID signatures respectively. We selected the threshold that maximized balanced accuracy when predicting MSI-PCR labels, just using the per-sample number of SBS and ID. Samples below these thresholds were designated a 0 activity for the corresponding mutational signatures. For the tumour–normal WES training data, these minima were 563 SBS and 19 ID.

Because this threshold is a property of the assay, not the tumour, we recomputed it for each of the sequencing panels assessed in this study. To do so, we restricted the SBS and ID in the training set to ones captured by the concerned targeted sequencing panel and re-assess the threshold optimization. Therefore, a technology specific threshold was established (Supplementary Tables 2 and 3). These in turn were used as the so-called indel and sbs rate rules. A sample is called MMRd when its per-sample indel (or SBS) count reaches the threshold of its panel.

Although this method could be performed to select a SBS and ID threshold for all targeted sequencing panels independently of their size, in ultra small panels accuracy can drop significantly. To account for this, we evaluated which was the minimum number of SBS and ID that independently of the sequencing technology could result in an accurate MMRd diagnosis. Therefore, using the training set we iteratively selected a specific number of SBS and ID mutations and evaluated the diagnosis using a balanced-class weight, unregularized logistic regression classifier trained just with SBS or ID signatures. To perform this analysis 100 random subsamples of the catalogue at each mutation count were performed. Then, signatures were refitted on each subsample per sample and MMR prediction was evaluated.

Whenever the median of the percentage of agreement across samples reached 90%, the indel threshold was set. This showed that 3 indels was the lowest to reliably predict MMR status using a 90% agreement with the MMR status prediction using each sample full indels set. In contrast, 44 sbs were needed to reliably predict MMRd diagnosis using the 90% agreement criteria. For substitutions however, this procedure was performed only in the MMRd/MSI sample subset instead of the full set as for indels.

Using the indel ratio per exonic megabase in the TCGA, we assessed the minimum exonic capture length for a panel depending on the cancer type to have a reliable MMRd prediction using captureMMRd. To assess this, we obtained the number of indel and substitutions in the stomach and colorectal TCGA testing subsets as well as in the endometrial TCGA cohort and divided these number by 35.8Mb, which was taken as the exonic territory of the TCGA exome. Then we selected the 10th percentile of the MSI samples to obtain a conservative per-Mb indel rate.

This was applied to a subset of clinically widely deployed targeted panels to check captureMMRd suitability. Protein-coding intervals used as TMB denominators were taken from GENCODE v49^93^ and intersected with the panel target intervals.

#### Feature selection

Model development and feature selection were performed in Python v3.12.13 using scikit-learn v1.6.1. Feature selection was conducted exclusively within the training set using recursive feature elimination with cross-validation (RFECV), with a logistic regression estimator using the default parameters. The candidate set comprised eight variables available for every training sample: the summed MMRd-associated ID activity, the summed MMRd-associated SBS activity, the MSIsensor score, tumour mutational burden, the presence of a loss-of-function alteration in an MMR gene, the aneuploidy score, and the per-sample indel and SBS catalogue sizes. Candidate feature subsets were evaluated using a custom Bayesian information criterion (BIC)

Feature selection was repeated over 500 resamples of the training set (stratified 5-fold cross-validation repeated 100 times under a fixed random seed). At each iteration, features were eliminated sequentially, with a minimum of 1 retained feature. To quantify how reproducible the selected set was, recursive feature elimination was refitted to the optimal model size on every resample and the proportion of resamples retaining each candidate is reported in Fig. 1b; for this stability analysis the candidates were standardized within each training fold, so that the elimination order reflects standardized effect size rather than the units a feature happens to be expressed in. A model consisting of just the ID signatures (Sum of ID1, ID2 and ID7) associated with MMRd was the one maximizing the BIC-based score across cross-validation folds. Because both signature activities are obtained from the same signature-fitting step, the SBS activity was additionally assessed against the ID-only model over the same resamples: the two models returned identical calls in 477 of 500 resamples and differed by less than 0.1 percentage points in cross-validated balanced accuracy, and both features were therefore retained in the final classifier.

#### Logistic regression classifier

The final model was an unregularized logistic regression classifier implemented in scikit-learn v1.6.1^94^. Balanced class weight and default settings for other parameters were used. The classifier estimated the probability of MMRd from SBS and ID signatures. Hence, the final model resulted in a coefficient of 7.03 for the sum of ID signatures associated with MMRd, 15.44 for the sum of SBS signatures associated with MMRd and an intercept of −3.9214564. Of note, if a sample’s indels or substitutions fall below its assay threshold, the concerned signature is assigned an activity of zero.

The final model was fitted to the complete training set and evaluated without further fitting or threshold optimization on the held-out 30% of the STAD and COADREAD cohorts. Generalizability to validation cohorts was evaluated separately on data totally excluded from the model training or feature selection.

#### Performance assessment

A prespecified probability threshold of 0.5 was used to classify samples as MMRd. Model performance was evaluated against the available orthogonal reference standard: MSI-PCR or MMR-IHC depending on the cohort (Supplementary Table 1). Sensitivity and specificity were calculated separately for each cohort. Corresponding 95% confidence intervals were estimated using the Wilson score method for binomial proportions.

### Statistical analysis

Statistical analyses were performed in Python using SciPy and statsmodels. Sensitivity and specificity were reported as binomial proportions with 95% Wilson score confidence intervals (statsmodels proportion_confint, method ‘wilson’). Comparisons between independent groups of samples, such as FFPE against fresh-frozen specimens, were tested with two-sided Fisher’s exact test, and differences in sensitivity and specificity between groups were reported with 95% Newcombe confidence intervals (statsmodels confint_proportions_2indep, method ‘newcomb’). Statistical significance was considered when p<0.05.

## Author contributions

G.M. conceived and designed the study. D.G.-L., B.H. and G.M. developed the methodology and software of the study. B.C.-U., M.E.-R., A.C. and M.J.G. designed and performed next-generation sequencing, MSI-PCR and MMR-IHC experiments. D.G.-L., B.H., B.C.-U., M.E.-R., D.G.-S., A.C., A.G., M.M., J.F.G., D.H., A.R., M.J.G. and G.M provided access to data and/or contributed to the data collection, processing, and curation; E.C., G.T. and A.B. performed the histopathological review of the OVAseq cohort, including MMRd assessment by IHC. D.G.-L., M.J.G. and G.M. wrote the paper. M.J.G. and G.M. supervised the project. All authors had access to all of the data in the study. All authors approved the paper before submission.

## Acknowledgments

This work was supported by grants to G.M. from the Spanish Ministry of Science and Innovation (Grants PID2019-111356RA-I00 and PID2022-137042OB-I00 funded by MICIU/AEI/10.13039/501100011033 and by FEDER/UE). The Macintyre group is hosted by the Centro Nacional de Investigaciones Oncológicas (CNIO), which is supported by the Instituto de Salud Carlos III and recognized as a ‘Severo Ochoa’ Centre of Excellence (ref. CEX2024-001442-S). B.H. is supported by a grant from ‘La Caixa’ Foundation (ID 100010434; LCF/BQ/PR23/11980033). M.E-R. received the support of a fellowship from the Spanish Ministry of Science and Innovation (grant no. PRE2020-092155 funded by MICIU/AEI/10.13039/501100011033 and by FSE+). B.C-U. has received a doctoral ‘La Caixa’ INPhINIT retaining fellowship (ID 100010434; LCF/BQ/DR23/12000023). MJG is supported by the Ministerio de Ciencia, Innovación y Universidades (MICIU)/AEI/10.13039/501100011033 and ERDF, EU (Project PID2023-151298OB-I00). The results shown here are in whole or part based upon data generated by the TCGA Research Network: https://www.cancer.gov/tcga. The authors acknowledge the American Association for Cancer Research (AACR) for its financial and material support of the AACR Project GENIE registry and the participating consortium institutions for their commitment to data sharing. The interpretations presented are solely the responsibility of the authors. Data were obtained from the AACR Project GENIE BioPharma Collaborative colorectal cancer cohort, release CRC v2.0-public (released November 2022). Data used in this publication were generated by the National Cancer Institute Clinical Proteomic Tumor Analysis Consortium (CPTAC). The colorectal cohort analysed here (cBioPortal study coad_cptac_2019) was generated by CPTAC and is reported in the Vasaikar et al. 2019 reference already cited. The authors also acknowledge Memorial Sloan Kettering Cancer Center (MSK) and the investigators responsible for generating and publicly sharing the MSK-IMPACT, MSK-CHORD, and MSK circulating tumour DNA sequencing datasets. We also acknowledge the patients for their participation and the IdiPAZ Biobank (PT23-00028), integrated into the ISCIII Biobanks and Biomodels Platform, and the BioBank of the Fundación MD Anderson Cancer Center España - Hospiten (B.0000745), integrated into the Spanish National Biobanks Network, for their collaboration and support. We acknowledge Dra. Gema Moreno-Bueno (Universidad Autónoma de Madrid-Institute for Biomedical Research Sols-Morreale (IIBM), Spanish National Research Council & MD Anderson Cancer Center Madrid– Hospiten) for facilitating access to tumour samples and for her support throughout this work. Finally, the authors acknowledge the cBioPortal for Cancer Genomics team and all contributing investigators, institutions, and patients for making cancer genomic and clinical data accessible to the research community.

## Data availability

Publicly available genomic and clinical data analysed in this study were obtained from cBioPortal for Cancer Genomics (https://www.cbioportal.org/), including The Cancer Genome Atlas (TCGA), MSK-IMPACT, and additional published sequencing cohorts. Data from the AACR Project GENIE BioPharma Collaborative colorectal cancer cohort (BPC CRC v2.0-public) were obtained through the AACR Project GENIE cBioPortal (https://genie.cbioportal.org/). The individual studies, accession identifiers, and corresponding references are detailed in Supplementary Table 1. Institutional sequencing data from the OVAseq cohort will be made available under controlled access upon publication.

